# Atrial Fibrillation Ablation Outcomes by Hospital Academic Status

**DOI:** 10.1101/2025.10.05.25337365

**Authors:** Muhammad Raffey Shabbir, Khubaib Ahmad, Mahrukh Imtiaz, Ahmad Baig, Saeed Aftab Khan, Qasim Bashir, Meenal Sikander, Muhammad Osama

## Abstract

**Background:** Atrial fibrillation (Afib) is the most prevalent arrhythmia worldwide and catheter ablation has been established as an effective treatment modality. The outcomes can vary based on medical expertise and location of the procedure. This study evaluates the differences in outcomes of catheter ablation performed at non-academic compared to academic institutions.

**Methods:** A retrospective cohort study was conducted using the TriNetX US Collaborative Network. Adults (35–90 years), who underwent Afib ablation between Jan 1, 2010, to Jan 1, 2020, were included. Those with congenital malformations of circulatory system, rheumatic heart disease, ischemic cardiomyopathy, or prior myocardial infarction (MI) were excluded.Groups were stratified by hospital academic status and balanced using 1:1 propensity score matching. Outcomes were assessed within 365 days post-ablation. Patients with outcome prior to the time window were excluded and odds ratio was used for statistical comparisons with significance set at p<0.05.

**Results:** Following propensity score matching, the analysis revealed that patients undergoing atrial fibrillation ablation at non-academic institutions had significantly higher odds of requiring additional or redo ablation (OR: 1.844; 95% CI: 1.409–2.415) and developing acute kidney injury (OR: 1.534; 95% CI: 1.054–2.232) compared to those treated at academic institutions. Other post-ablation complications, including cardiac arrest (OR: 1.101; 95% CI: 0.466–2.599), cardiac tamponade (OR: 1.101; 95% CI: 0.466–2.599), esophageal perforation (OR: 1.000; 95% CI: 0.415–2.409), and hemorrhages or hematomas (OR: 0.909; 95% CI: 0.385–2.145), did not differ significantly between the two groups.

**Conclusion:** Catheter ablation of atrial fibrillation performed at academic hospitals resulted in better outcomes, potentially reflecting advanced technical expertise, post-op care and better institutional resources. These results highlight the importance of standardization of care and the need for increased access of high-standard care across healthcare settings. Future studies should investigate modifiable institutional factors and patient level variables driving this disparity.

## Introduction

Atrial fibrillation (AF) is the most common arrhythmia encountered in clinical practice, with an estimated global prevalence of 52.5 million individuals [1]. In the United States, the prevalence of AF is estimated to be around 3-5 million with a predicted increase to over 8 million by the next two decades[2]. Current treatment strategies for AF include anticoagulation based on stroke risk, rate and rhythm control, with anti-arrhythmic medications or catheter ablation [3]. Catheter ablation is the most common cardiac ablation procedure performed globally. It has been indicated for patients with AF in whom initial treatments have failed[3].

Long term outcomes of catheter ablation in AF include a significant decrease in mortality rates, heart failure hospitalizations and stroke incidence, and its comparative efficacy and benefits against traditional antiarrhythmic therapy alone are currently under debate[4]. Post procedural success of the treatment has increased significantly over the recent years. An analysis conducted by Perino et al. estimated the success rate to have increased from 73.1% in 2003 to 77.1% in 2016, with an average increase of 1.6% when adjusting for confounding data [5]. Similar rates of success have reported in geriatric patient population with AF, highlighting its importance as a treatment modality for arrhythmias unresponsive to medication [6]. The Heart Rhythm Society (HRS) and American College of Cardiology has proposed a ‘same-day discharge’ for intra-cardiac catheter ablation procedures to reduce hospitalization costs and healthcare utilization, based on the high success rate and low complication risk of the procedure [7]. This proposal was based on the results of several published studies and trials with heterogeneity in terms of access to specialists, and conduction in a high volume setups such as tertiary or academic setting 8, 9]

Existing data, when subjected to further analysis, reveals lower success and higher complication rates in catheter ablation conducted in low volume settings, revealing the non-generalizability of published outcomes across all settings [10]. Procedural disparities across various clinical settings play a significant role in post-ablation complications [11]. Studies that are conducted in controlled academic environments usually enroll a large number of participants, have a large-scale access to specialists, and follow latest treatment protocols and guidelines, whereas nonacademic settings lack sufficient patient load, and have no surgical cardiothoracic, electrophysiology departments or rapid response teams [12-14].

Considering the facts, the current study aims to delineate differences in mortality in catheter ablations conducted in academic versus those conducted in nonacademic settings.

## Methods

### 2.1. IRB Approval

This retrospective cohort study does not require Institutional Review Board (IRB) approval because it complies with the Strengthening the Reporting of Observational Studies in Epidemiology (STROBE) guidelines and does not involve human participant research [15].

### 2.2. Data source

The TriNetX (TriNetX, USA) platform was used in this study to acquire the relevant data for analysis. TriNetX is a global federated health research network that provides access to de-identified electronic medical records (EMRs), including diagnoses, procedures, medications, laboratory values, and genomic information, across numerous medical and large healthcare organizations (HCOs), which helps in conducting a comprehensive evaluation of actual clinical outcomes among patients. Additional information pertaining to this database can be found elsewhere in the literature [16]

### 2.3. Patient cohort

We retrieved data from health care organizations that are a part of TriNetX’ s US Collaborative Network from January 1, 2010 to January 1, 2020 of adults aged 35 to 90 years with a diagnosis of atrial fibrillation, which was identified by using the International Classification of Diseases, Tenth Revision (ICD-10) code I48, who underwent catheter ablation for AF at any time in their medical record. The catheter ablation procedure was identified using the Unified Medical Language System (UMLS) Current Procedural Terminology (CPT) 93656 or 93657. Two individual patient cohorts were formed based on the academic status of the health care facility, non-academic and academic (Figure 1). Cohort 1 consisted of patients who underwent ablation in non-academic center/facility and named as “Non-Academic Group”, whereas cohort 2 consisted of patients who underwent ablation in academic center/facility and named as “Academic Group”. For each patient, the index date was defined as the first record of an AF ablation procedure. The follow-up extended until the end of each patient’s available electronic record or till 14^th^ May 2025.

**Fig 1:**
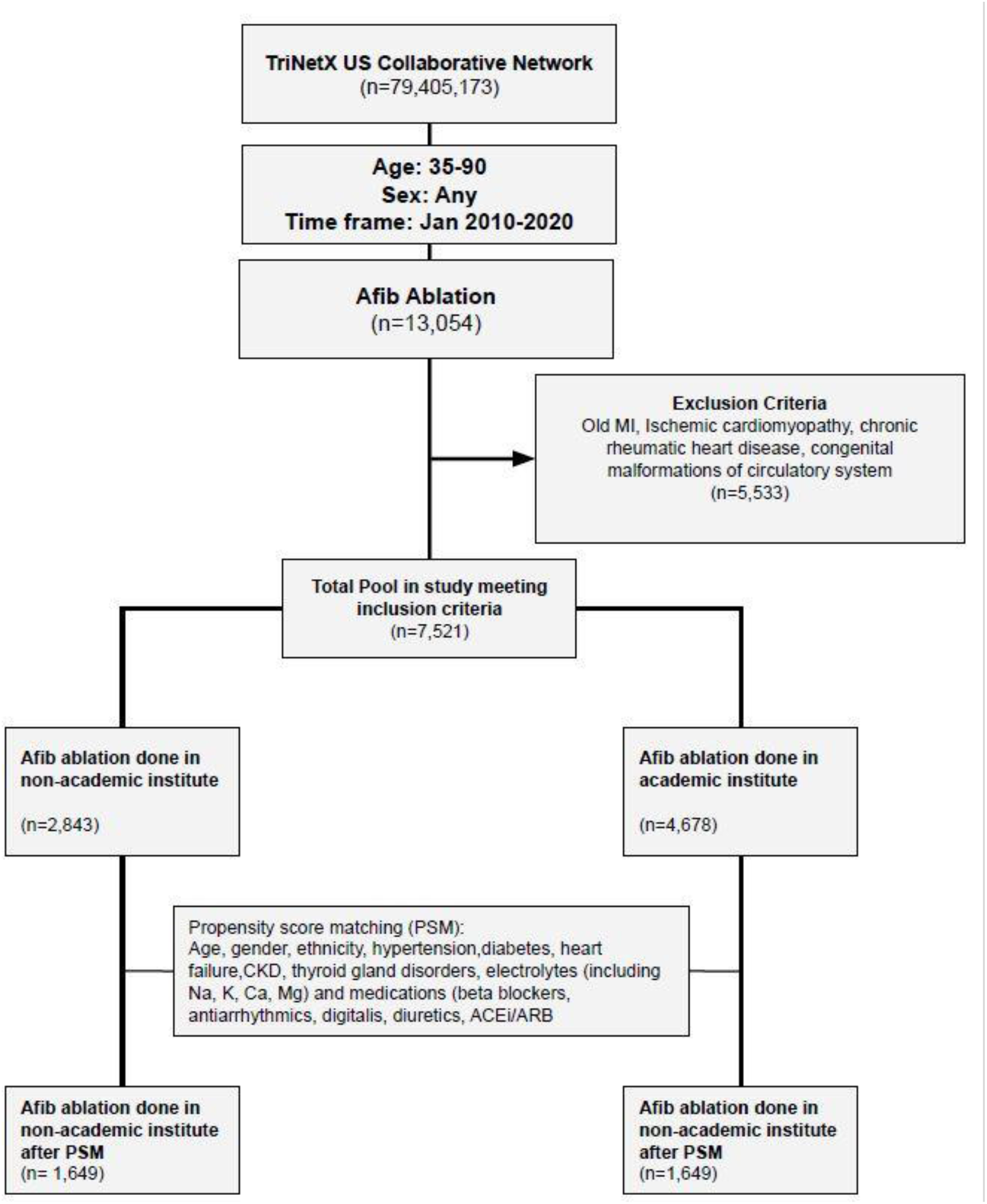
STROBE flow chart. US: United States, MI: Myocardial Infarction, CKD: Chronic Kidney Disease

### 2.4. Propensity score matching (PSM)

Propensity score matching (PSM) was used to create a balanced comparison between the two cohorts, allowing for the matching of patients based on key demographic and clinical factors to minimize confounding (Table 1). These included baseline characteristics such as age, sex, race or ethnicity, body mass index (BMI), hypertension, diabetes, disorders of lipoprotein metabolism and other lipidemia, heart failure (HF), chronic kidney disease (CKD), chronic obstructive pulmonary disease (COPD), thyroid gland disorders, and concurrent cardiovascular (CV) medications (beta-blockers, anti-arrhythmic, diuretics, ACEi, Digitalis Glycosides). Additionally, laboratory reports, such as serum electrolytes (sodium, potassium, calcium, magnesium) were also matched (Table 1). After completing the matching process, we calculated a standardized difference to evaluate the balance of patient characteristics between the matched cohorts. A standardized difference <0.1 indicates that the groups are well-balanced (Figure 2).

**Table 1:**
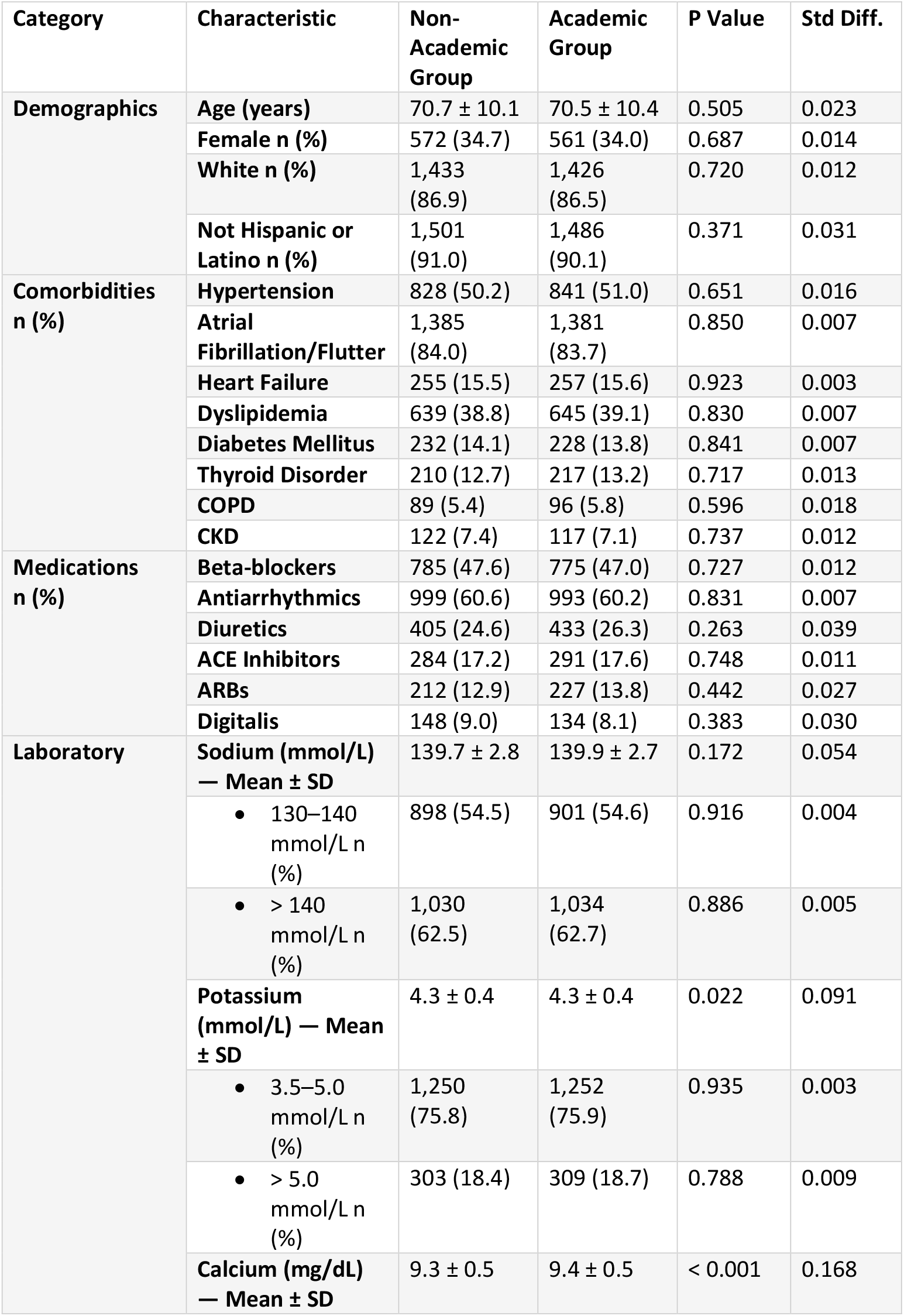
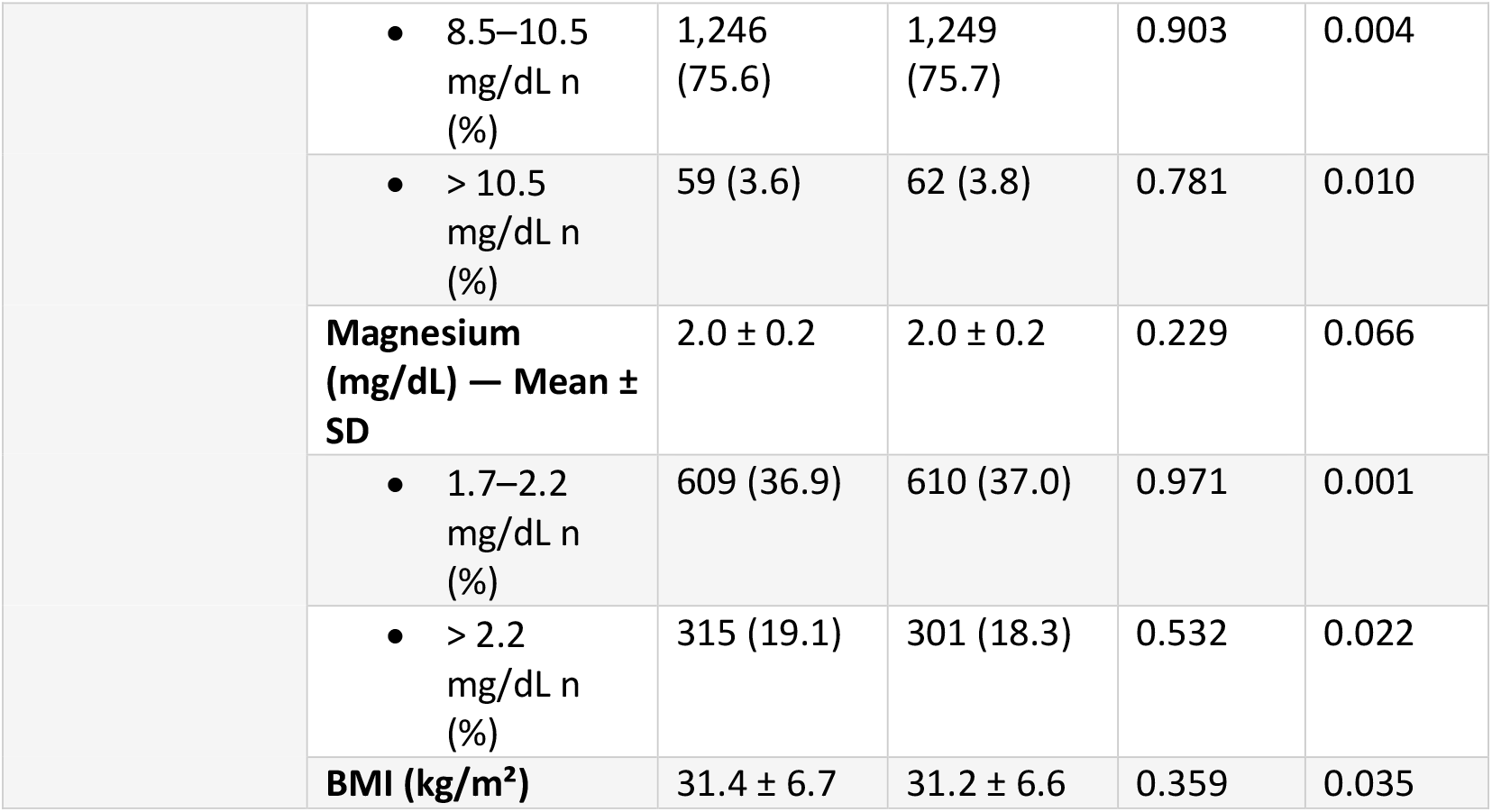
Baseline Characteristics (After Propensity Score Matching, N = 1,649 per Group). Numerical data is represented as mean ± standard deviation, while categorical data is expressed as count (%). Abbreviations: SD: Standard Deviation, Std diff: Standard Difference, BMI: Body mass index.

| Category | Characteristic | Non-Academic Group | Academic Group | P Value | Std Diff. |
| --- | --- | --- | --- | --- | --- |
| Demographics | Age (years) | 70.7 ± 10.1 | 70.5 ± 10.4 | 0.505 | 0.023 |
|  | Female n (%) | 572 (34.7) | 561 (34.0) | 0.687 | 0.014 |
|  | White n (%) | 1,433 (86.9) | 1,426 (86.5) | 0.720 | 0.012 |
|  | Not Hispanic or Latino n (%) | 1,501 (91.0) | 1,486 (90.1) | 0.371 | 0.031 |
| Comorbidities n (%) | Hypertension | 828 (50.2) | 841 (51.0) | 0.651 | 0.016 |
|  | Atrial Fibrillation/Flutter | 1,385 (84.0) | 1,381 (83.7) | 0.850 | 0.007 |
|  | Heart Failure | 255 (15.5) | 257 (15.6) | 0.923 | 0.003 |
|  | Dyslipidemia | 639 (38.8) | 645 (39.1) | 0.830 | 0.007 |
|  | Diabetes Mellitus | 232 (14.1) | 228 (13.8) | 0.841 | 0.007 |
|  | Thyroid Disorder | 210 (12.7) | 217 (13.2) | 0.717 | 0.013 |
|  | COPD | 89 (5.4) | 96 (5.8) | 0.596 | 0.018 |
|  | CKD | 122 (7.4) | 117 (7.1) | 0.737 | 0.012 |
| Medications n (%) | Beta-blockers | 785 (47.6) | 775 (47.0) | 0.727 | 0.012 |
|  | Antiarrhythmics | 999 (60.6) | 993 (60.2) | 0.831 | 0.007 |
|  | Diuretics | 405 (24.6) | 433 (26.3) | 0.263 | 0.039 |
|  | ACE Inhibitors | 284 (17.2) | 291 (17.6) | 0.748 | 0.011 |
|  | ARBs | 212 (12.9) | 227 (13.8) | 0.442 | 0.027 |
|  | Digitalis | 148 (9.0) | 134 (8.1) | 0.383 | 0.030 |
| Laboratory | Sodium (mmol/L) — Mean ± SD | 139.7 ± 2.8 | 139.9 ± 2.7 | 0.172 | 0.054 |
|  | • 130–140 mmol/L n (%) | 898 (54.5) | 901 (54.6) | 0.916 | 0.004 |
|  | • > 140 mmol/L n (%) | 1,030 (62.5) | 1,034 (62.7) | 0.886 | 0.005 |
|  | Potassium (mmol/L) — Mean ± SD | 4.3 ± 0.4 | 4.3 ± 0.4 | 0.022 | 0.091 |
|  | • 3.5–5.0 mmol/L n (%) | 1,250 (75.8) | 1,252 (75.9) | 0.935 | 0.003 |
|  | • > 5.0 mmol/L n (%) | 303 (18.4) | 309 (18.7) | 0.788 | 0.009 |
|  | Calcium (mg/dL) — Mean ± SD | 9.3 ± 0.5 | 9.4 ± 0.5 | < 0.001 | 0.168 |
|  | 8.5–10.5 mg/dL n (%) | 1,246 (75.6) | 1,249 (75.7) | 0.903 | 0.004 |
|  | > 10.5 mg/dL n (%) | 59 (3.6) | 62 (3.8) | 0.781 | 0.010 |
|  | **Magnesium (mg/dL) — Mean ± SD** | 2.0 ± 0.2 | 2.0 ± 0.2 | 0.229 | 0.066 |
|  | 1.7–2.2 mg/dL n (%) | 609 (36.9) | 610 (37.0) | 0.971 | 0.001 |
|  | > 2.2 mg/dL n (%) | 315 (19.1) | 301 (18.3) | 0.532 | 0.022 |
|  | **BMI (kg/m<sup>2</sup>)** | 31.4 ± 6.7 | 31.2 ± 6.6 | 0.359 | 0.035 |

**Figure 2.**
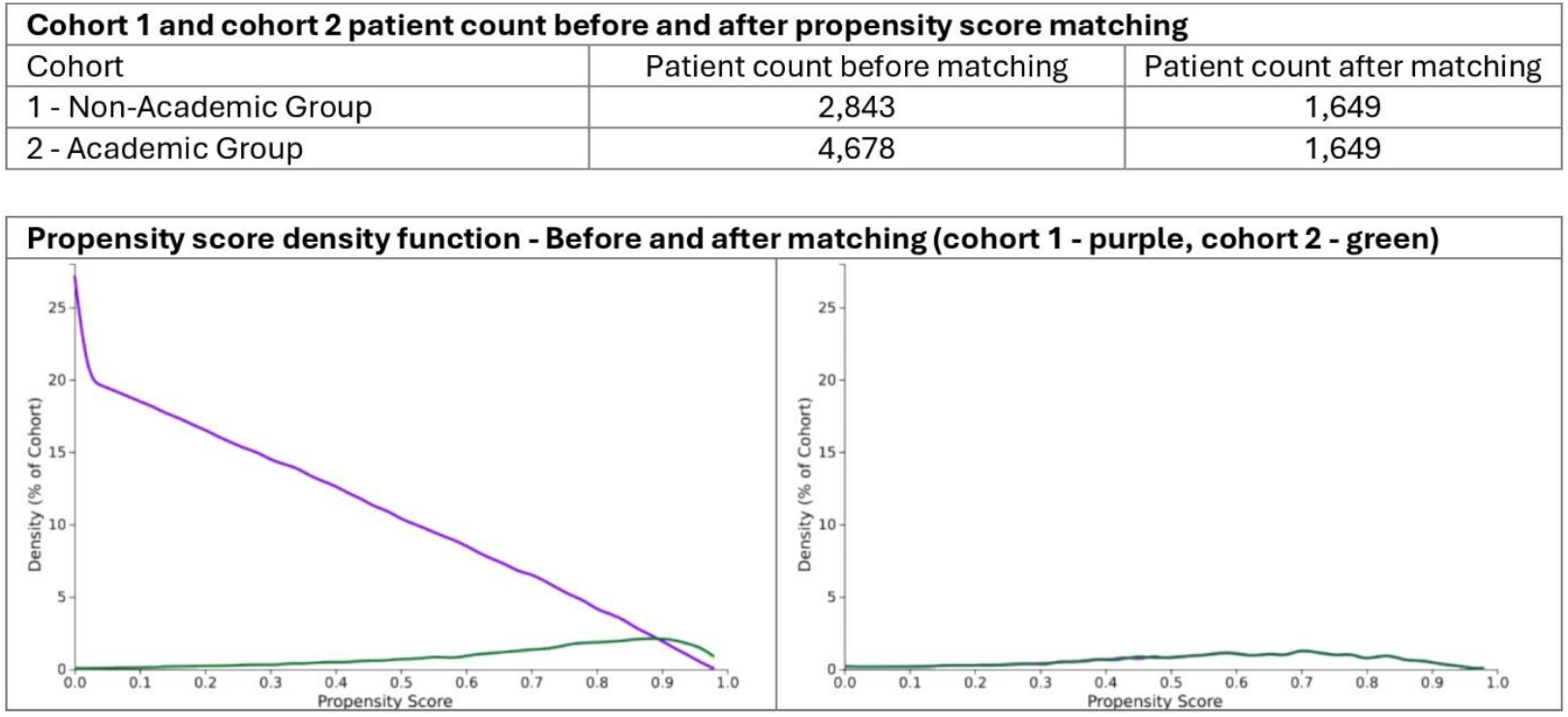
The graph demonstrates the propensity score before and after matching cohort 1 – purple and cohort −2 green. The curves on the right show imbalance between both the cohorts but after matching the curves nearly are overlapping as shown on the right confirming that the cohorts were well balanced and comparable for analysis.

### 2.5. Outcomes

The outcomes assessed in this study included additional or re-do ablation procedures (CPT 93657 & 93656), cardiac tamponade (ICD: I31.4), esophageal perforation (ICD: K22.3), hemorrhages/hematomas (ICD: I97.6 and I97.410), acute kidney injury (ICD: N17), cardiac arrest (ICD: I46) (Table 2, Figure 3).

**Table 2.** Odds ratio, risk ratio, and risk difference for post-ablation outcomes of atrial fibrillation in non-academic vs. academic institutions after propensity score matching. Abbreviations/key: n/N Number of patients experiencing the outcome/Total patients in the group; CI=Confidence Interval; RD=Risk Difference; RR=Risk Ratio; OR=Odds ratio

| Outcomes | n/N (Risk %) |  | Risk Difference<br>RD (95% CI) | Odds Ratio<br>OR (95% CI) | Risk Ratio<br>RR (95% CI) |
| --- | --- | --- | --- | --- | --- |
|  | Non-Academic<br>Institutions | Academic<br>Institutions |  |  |  |
| **Additional/Redo Ablation** | 157/1649 (9.5) | 89/1649 (5.4) | 0.041 (0.023, 0.059) | 1.844 (1.409, 2.415) * | 1.764 (1.373, 2.267) |
| **Acute Kidney Injury (AKI)** | 71/1649 (4.3) | 47/1649 (2.9) | 0.015 (0.002, 0.027) | 1.534 (1.054, 2.232) * | 1.511 (1.052, 2.170) |
| **Cardiac Arrest** | 11/1649 (0.7) | 10/1649 (0.6) | 0.001 (-0.005, 0.006) | 1.101 (0.466, 2.599) | 1.1 (0.468, 2.583) |
| **Cardiac Tamponade** | 11/1649 (0.7) | 10/1649 (0.6) | 0.001 (-0.005, 0.006) | 1.101 (0.466, 2.599) | 1.1 (0.468, 2.583) |
| **Esophageal Perforation** | 10/1649 (0.6) | 10/1649 (0.6) | 0 (-0.005, 0.005) | 1 (0.415, 2.409) | 1 (0.417, 2.396) |
| **Hemorrhages/Hematomas** | 10/1649 (0.6) | 11/1649 (0.7) | -0.001 (-0.006, 0.005) | 0.909 (0.385, 2.145) | 0.909 (0.387, 2.135) |

\* $p < 0.05$

**Figure 3:**
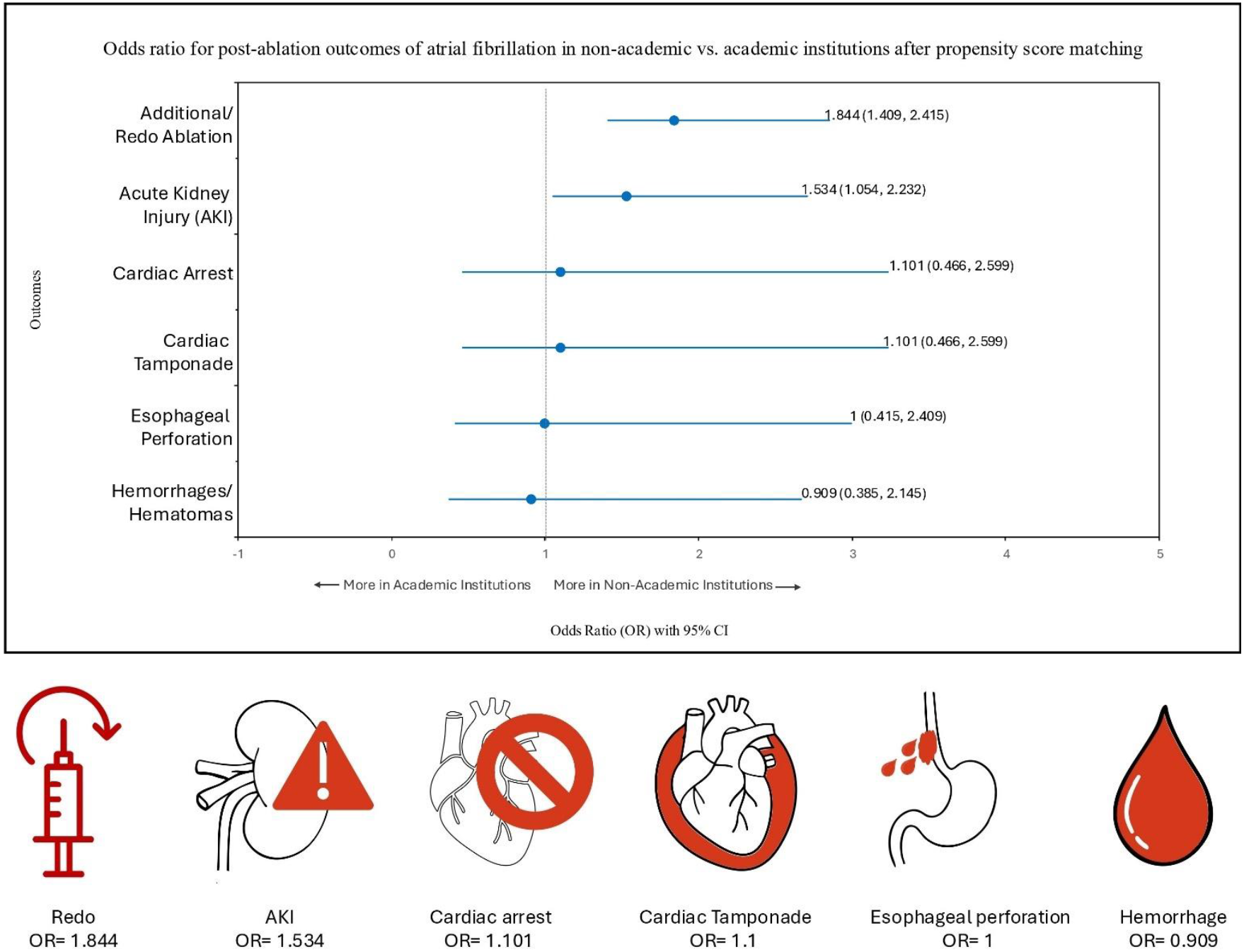
Forest plot with illustration: Odds ratio with 95% CI for post-ablation outcomes of atrial fibrillation comparing non-academic vs. academic institutions after propensity score matching. The results show that the odds of requiring additional/redo ablation and developing acute kidney injury were significantly higher in the non-academic centers as compared to the academic centers. All other complications such as cardiac arrest, cardiac tamponade, esophageal perforation, hemorrhages/hematomas though higher in the non-academic institutions show no significant difference between academic and non-academic institutions.

### 2.6. Statistical analysis

All analyses were executed using the TriNetX platform integrated tool, accessed on May 14, 2025, along with the utilization of R survival package version 3.2-3. The chi-square test was used to compare the categorical variables between the two cohorts for the assessment of baseline characteristics. The t-test was employed for the assessment of continuous variables. Risk analyses yielded odds ratio (OR), absolute risk, risk difference (RD), and risk ratio (RR). Categorical variables were compared by chi-square test; continuous variables by two-sample t-tests. A two-sided P < 0.05 indicated statistical significance.

## Results

A total of 7521 adults underwent atrial fibrillation ablation between January 1, 2010, and January 1, 2020, who were identified from the TriNetX US Collaborative Network. Patients with congenital circulatory malformations, rheumatic heart disease, ischemic cardiomyopathy, or a history of myocardial infarction were excluded from the study. The population was divided based on the type of hospital where the ablation occurred, with 2,843 patients treated at non-academic institutions and 4,678 patients treated at academic institutions. To address baseline imbalances, 1:1 propensity score matching was performed, resulting in matched cohorts of 1,649 patients in each group (Table 1, Figure 2). After the matching process, the baseline demographics, comorbidities, and treatment characteristics were well balanced between the two groups, as indicated by standardized mean differences of less than 0.1. The clinical outcomes after the procedure were assessed within 365 days using odds ratios (ORs) with 95% confidence intervals (CIs) to compare post-procedural complication rates. A p-value of <0.05 was considered statistically significant.

Post-ablation outcomes of atrial fibrillation from both academic and non-academic institutions are presented with Risk Difference, Odds Ratio, and Risk Ratio (Table 2, Figure 3,4). A total of 157 patients who were treated at non-academic institutions required a repeat ablation, compared to 89 patients who received treatment at academic centers. The odds of needing a repeat ablation were 1.844 times higher for patients at non-academic institutions than for those at academic centers (95% CI 1.409–2.415, p< 0.05). The odds of Acute Kidney Injury were 1.534 times higher in patients treated at non-academic institutions, with 71 cases compared to 47 at academic institutions. This difference was statistically significant (95% CI 1.054–2.232, p < 0.05) *p<0.05

**Figure 4:**
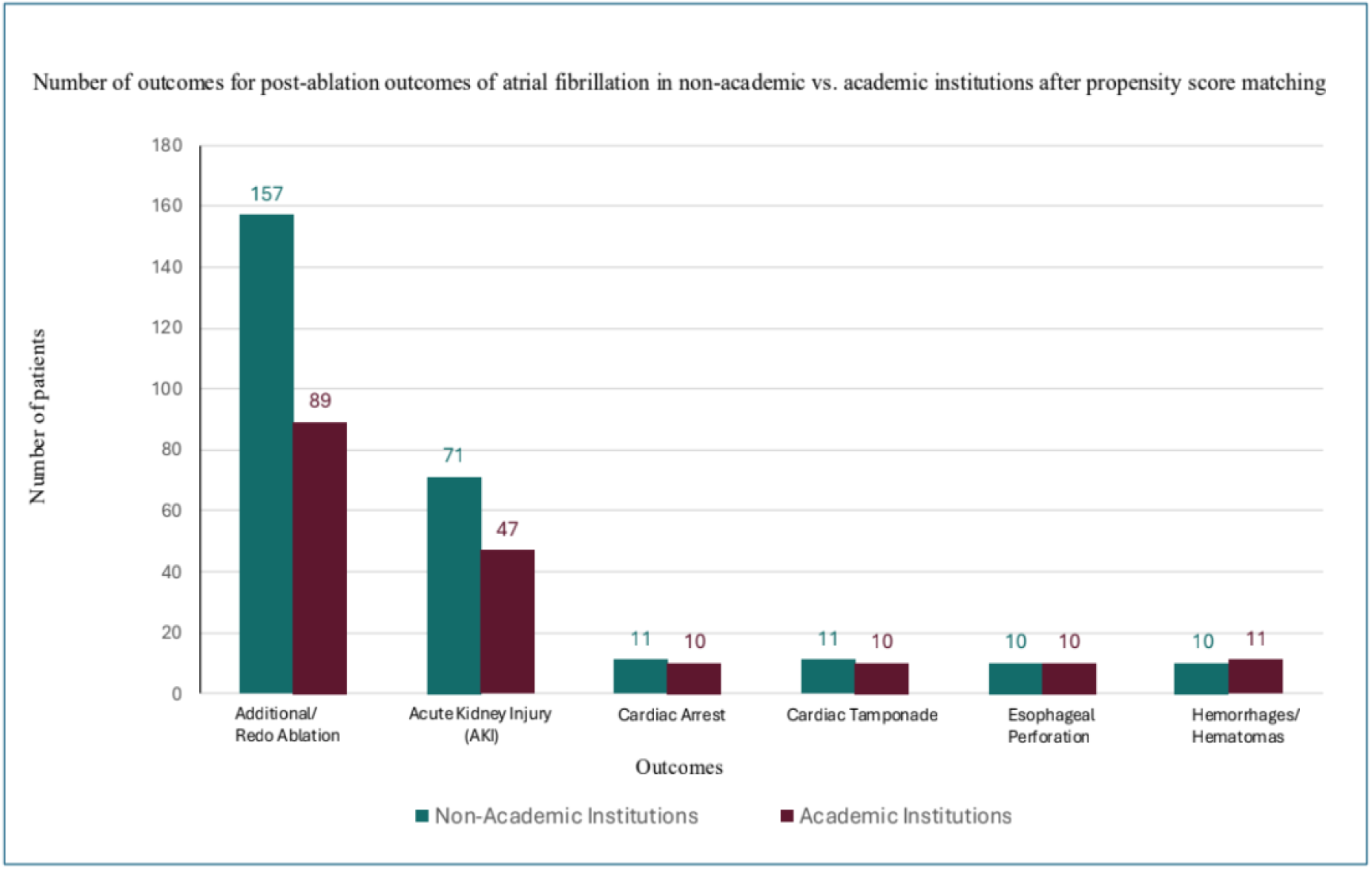
Bar chart comparing the number of post-ablation outcomes between academic and non-academic institutions shows that redo/ additional ablation occurs more frequently in non-academic centers as compared to the academic centers while other complications such as cardiac arrest, cardiac tamponade, esophageal perforation and hemorrhage/hematoma are almost similar between both groups.

No statistically significant differences were observed in the incidence of other complications, including cardiac arrest (OR=1.101, 95%CI 0.466–2.599), cardiac tamponade (OR=1.101, 95%CI 0.466–2.599), esophageal perforation (OR=1.000, 95%CI 0.415–2.409), or hemorrhages/hematomas (OR=0.909, 95%CI 0.385–2.145).

Patients undergoing AF ablation in non-academic institutions had a significantly higher likelihood of requiring repeat ablation and developing acute kidney injury. No significant differences were observed in life-threatening complications such as cardiac arrest, tamponade, esophageal injury, or bleeding. (Table 2).

## Discussion

Our study aimed to investigate clinical outcomes following atrial fibrillation (AF) catheter ablation between academic and non-academic institutions. Our findings demonstrate that patients undergoing ablation in non-academic institutions were more likely to require repeat ablation and develop acute kidney injury (AKI) within one year of the procedure compared to patients treated at academic institutions. However, no significant differences were observed for other life-threatening complications, including cardiac arrest, tamponade, esophageal perforation, or hemorrhage/hematoma.

Despite the increased disease severity and complexity of patients treated at academic hospitals, prior literature has shown improved mortality outcomes for common medical conditions in patients treated at these institutions [17-20]. These differences have also been reported in common surgical procedures; however, data for advanced procedures is limited. In a large-scale analysis by Ngo et al., substantial variability in 30-day post-ablation complications was reported across institutions, supporting the notion that outcomes differ by institutional quality and resource availability [11] Deshmukh et al., utilizing data from over 93,000 ablations in the United States, reported that teaching hospitals demonstrate lower rates of in-hospital complications compared to non-teaching hospitals, likely driven by higher procedural volumes and availability of advanced cardiac services [13]. Despite similar patient characteristics, our study showed higher repeat ablation rates in non-academic institutions during a one-year follow-up. This discrepancy suggests that institutional factors may play a role, such as less patient follow-up support, fewer experienced electrophysiologists, and limited access to advanced mapping or imaging technologies. Di Biase et al. noted in a multicenter study that academic institutions tend to have real-time imaging and advanced mapping tools that reduce the likelihood of ablation failure [12].

We hypothesize that the higher likelihood of acute kidney injury (AKI) observed in non-academic institutions reflects modifiable, system-level differences in peri-operative care and procedural efficiency. First, protocolized hydration is one of the most effective strategies for preventing contrast-associated AKI (CA-AKI), yet implementation is inconsistent outside of academic centers. Randomized trials have demonstrated that isotonic intravenous hydration significantly reduces the incidence of CA-AKI compared with hypotonic regimens or no standardized protocol [21, 22]. Second, procedure duration and contrast stewardship are critical determinants of renal safety. Prolonged procedures are strongly correlated with higher contrast exposure, and exceeding the maximum allowable contrast dose (MACD) independently predicts CA-AKI across cardiovascular interventions (Nyman et al., 2019) [23].In AF ablation, advanced workflows available in many academic centers, including intracardiac echocardiography (ICE), electroanatomic mapping, and pressure-guided techniques, enable low-or even zero-contrast ablation while maintaining safety and efficacy, thereby reducing renal stress [24].

Finally, early nephrology involvement and overall institutional readiness are key for preventing AKI and other organ complications. Acute kidney injury, which was more prevalent in non-academic institutions in our study, may be attributed to less robust perioperative management, limited nephrology consultation, or prolonged procedure times. As highlighted by Piasecki et al., rapid response systems including nephrology and critical care backup are associated with reductions in non-cardiac complications such as acute renal failure (Piasecki et al., 2006) [14]. Collectively, these mechanisms, including hydration protocolization, contrast minimization through advanced imaging and mapping, timely nephrology involvement, and comprehensive institutional preparedness, provide a biologically and operationally plausible explanation for the higher AKI rates we observed in non-academic centers, despite similar baseline patient characteristics. These findings underscore the importance of system-level interventions, beyond patient factors alone, to reduce AKI risk and optimize outcomes across diverse procedural settings.

Some studies have shown similar outcomes across various institutional settings. Data from the National Inpatient Sample analyzed by Gehi et al. reported that no significant difference in post-ablation complications was found between high- and low-volume hospitals after adjustment for baseline characteristics [25]. One possible explanation for these discrepancies lies in methodological variation across studies. While our study utilized propensity score matching to balance baseline variables such as comorbidities and demographics, other studies relied on administrative data with potential limitations in database coding, confounder adjustment, and institutional misclassification. The lack of significant differences in operative complications such as tamponade, cardiac arrest, esophageal injury, and bleeding observed in our study is supported by reports from several multicenter evaluations. Benali et al. concluded that serious complications were infrequent and not significantly different across academic and non-academic settings, likely due to improving procedural safety technology across institutions [26]. This finding reinforces the idea that institutional disparities may primarily affect long-term outcomes and complications related to procedural planning and follow-up, rather than catastrophic events occurring during the ablation.

Our results support a nuanced view in which academic centers may serve not only as high-volume hubs but also offer comprehensive multidisciplinary care, rapid response teams, and adherence to the latest guidelines, potentially enhancing outcomes. Our study contributes to the growing literature suggesting that not all procedural settings are equal when managing complex cardiac arrhythmias and that institutional characteristics— particularly academic versus non-academic designation—may be important variables in risk stratification and procedural planning. While acute kidney injury and repeat ablation rates were higher in non-academic centers in the immediate follow-up period, it remains unclear if these events translate into worse long-term cardiovascular outcomes, including stroke or all-cause mortality. Future studies assessing composite endpoints over multi-year periods may help clarify this.

## Limitations

One limitation of our study is the potential for residual confounding despite rigorous matching. For instance, we could not assess operator-specific characteristics (e.g., years of experience) or hospital infrastructure details that may influence outcome disparities. Additionally, our analysis lacked granular data on peri-procedural practices such as contrast volume management, hydration protocols, and follow-up intensity, which may contribute to differences in AKI and repeat ablation rates. We were also unable to capture patient adherence to post-ablation care or outpatient monitoring, factors that could affect long-term outcomes. Finally, as with all observational studies, unmeasured socioeconomic or regional differences between academic and non-academic centers may have influenced both procedural planning and complication rates.

## Conclusion

Our study compared outcomes of atrial fibrillation ablation between academic and non-academic institutions and found that patients treated at non-academic centers experienced higher rates of repeat ablation and acute kidney injury (AKI) within one year, although rates of major peri-procedural complications such as tamponade or cardiac arrest were similar. These differences likely reflect institutional factors, including access to advanced imaging and mapping technologies, standardized hydration and contrast protocols, and multidisciplinary perioperative support more commonly available in academic hospitals. Our findings suggest that while procedural safety has improved across settings, disparities remain in long-term outcomes related to planning, follow-up, and resource availability.

Future studies are needed to determine whether these short-term differences translate into worse long-term cardiovascular events.

## Data Availability

All data generated or analyzed during this study are included in this article. Further inquiries can be directed to the corresponding author.

https://live.trinetx.com/

## Acknowledgments

None

